# MULTIPLE ENDEMIC EQUILIBRIA IN AN ENVIRONMENTALLY-TRANSMITTED DISEASE WITH THREE DISEASE STAGES

**DOI:** 10.1101/2024.03.21.24304681

**Authors:** José Manuel Islas, Ruth Corona-Moreno, Jorge X. Velasco-Hernández

**Affiliations:** Instituto de Matemáticas Unidad Juriquilla, Boulevard Universitario 3001, Juriquilla, 76230, Querétaro, México

**Keywords:** Threshold behavior, backward bifurcation, environmental transmitted disease, mathematical model

## Abstract

We construct, analyze and interpret a mathematical model for an environmental transmitted disease characterized for the existence of three disease stages, acute, severe and asymptomatic where severe and asymptomatic cases may present relapse between them. Transmission dynamics driven by the contact rates (as normally occur in directly-transmitted or vector-transmitted diseases) only occurs when a parameter *R*_*_ *>* 1. In this case, the forward transcritical bifurcation that exists for *R*_*_ *<* 1, becomes a backward bifurcation, producing multiple steady-states, a hysteresis effect and dependence on initial conditions. A threshold parameter for an epidemic outbreak, independent of *R*_*_ is only the ratio of the external contamination inflow shedding rate to the environmental clearance rate. *R*_*_ describes the strength of the transmission to infectious classes other than the *I*-(acute) type infections. The epidemic outbreak conditions and the structure of *R*_*_ appearing in this model are both resposible for the existence of endemic states.

## 1 Introduction

Environmentally-transmitted infectious diseases (ETID) constitute important health concerns around the globe for a large fraction of the world population. Diseases such as amoebiasis, cholera, toxoplasmosis, leptospirosis and many other pathogens are transmitted via contaminated water, soil and surfaces, and affect both animals and humans [2, 12, 15, 16, 34]. The relevance of study of these diseases lies in the variety of illness pathways that can develop in the host; for instance, they can be mild, severe or, even asymptomatic [2, 15, 17]. This variety of scenarios emergence from the existence of different strains able to infect a host, the tolerance level of susceptible individuals and the antimicrobial resistance that infectious agents can develop [8, 19, 28].

Diseases that are transmitted by exposure to contaminated environments such as air, water, and surfaces are of major public health importance documented by an ample bibliography [3, 11, 32]. An example of a bacteria that can present a variety of strains with different pathogenic qualities is *Vibrio cholera*. Strains of the serogroups O1 and O139 can cause the diarrheal disease; but *V. cholerae* bacteria belonging to other serogroups (designated non-O1, non-O139) are rather associated with gastrointestinal or extraintestinal infections [32, 27]. To treat these diseases, the use of antimicrobials is widespread with different levels of success [23, 24]. Antimicrobial use is a known selection factor that may generate bacterial or pathogen strains that are resistant to them thus diminishing the efficacy of population strategies for disease control and mitigation [31].

The factors that contribute to the development of different illness levels involve both the pathogenic strain involved and the immune response of the individual. This last factor is, likewise, subject to individual conditions of overall health such as nutritional status, host immunocompromised condition, and concomitant infections and comorbidities [28]. Continuous exposure to a contaminated environment may induce differential susceptibility in the host, tolerance to disease or resistance (immunity) to infection [6]. Likewise, the immune or susceptibility status of the host when exposed to infection may also determine the pathway of illness that may result. The use of antimicrobials against pathogens can alter or enhance either of these possibilities because they can select for strains associated with particular forms of illness [4, 36].

Pathogens, in particular bacterial pathogens, do coexist with our microbiome in our digestive, respiratory, urogenital tracts and skin; in many cases human hosts are simply their carriers [21]. This condition, however, may change and the asymptomatic carrier can develop disease that sometimes can be serious with those who develop it becoming at high risk of death or recurrent damaging morbidity [21, 22]. The environment is a complex set of conditions that determine or favor a mixture of pathogens subject to a variety of selection pressures amongst which antibiotics are particularly important. The microorganismic mixture that exists in a given environment (we can think on water bodies or hospitals to fix ideas), exposes hosts to a diversity of pathogens whose acquisition becomes a random event. In fact, it is known, in general, that a certain fraction of the population is exposed to environmental-transmitted pathogens that lead to a normal disease pathway and other fraction, usually much larger [21, 33], become asymptomatic carriers of microorganisms that can be resistant to antibiotic treatment. As individuals come into contact with a contaminated environment, prolonged exposure to antimicrobial resistance pathogens may weaken or significantly downgrade their overall health status making them more susceptible to severe disease [6, 20, 30].

For years, efforts have been made to implement environmental sanitation measures to eradicate environmental-transmitted diseases based on a variety of strategies and tools; however, there are regions or countries where, despite technological advances, sanitary resources are not sufficient or readily available to eradicate or, at least, mitigate the impact of these illnesses [7]. For instance, it is estimated that for cholera there are approximately 4 million cases and 143,000 deaths per year [9].

There are several approaches that have been developed to understand the dynamics of environmental transmitted diseases [5]. We use the pioneering model in [10] as the basic framework of our work, that has been shown to be appropriate and general enough. For instance, in [13] a between-within host mathematical model is proposed to study the dynamic of toxoplasmosis infection, an environmental transmitted disease; in [35], a model is proposed to analyze the dynamics of Giardia spread, a disease affecting both animals and humans. This model links two separate SI models, each representing either the animal or human population, and incorporates a compartment that accounts for the free-living life stage of the protozoan pathogen in water. The model assumes that disease transmission among animals occurs solely through direct contact, but there is also a contribution from contaminated water. When humans ingest contaminated water, they can become infected and further propagate the disease within their population. Other approaches based on partial differential equations models have been proposed to study the effect of spatial dynamics as in [18] where the propagation of fomites and mites that survive in the bedding chamber of wombat burrows is analyzed.

It is of interest, therefore, to study the consequences of having different susceptibility levels and disease pathways in the prevalence of environmental-transmitted diseases. All of these assumption are combined together on the present work. In §2 we construct and describe a mathematical model that is used to study the plausible dynamics of a general environmental-transmitted disease. Its steady-states and their stability analysis, depending on an environmental threshold parameter 𝒯_0_, are studied in §3. Subsequently, in §4, we show that this parameter controls the existence of a backward bifurcation that implies the coexistence of an stable disease-free equilibrium point and an stable endemic equilibrium point, when the inflow rate of external environmental contamination is lower than its clearance rate. Afterwards, in §5 simulations for the main theorem are shown as well as a numerical analysis for the general model showing general condition in which the backward bifurcation is maintained. The discussion and conclusions of this work are found in §5.1 and §6.

## 2 Mathematical model

In this section we present a mathematical model for a generic ETID. The framework is general and we are interested specifically on studying the interplay between three levels of illness (asymptomatic, acute and severe) and the induced dynamical behavior that their parameters control.

We model the dynamic of the spread of antimicrobial sensitive bacteria, that follows a susceptible - infectious - susceptible pattern, common to many bacterial diseases, competing only with an antimicrobial resistant (AMR) bacteria, that follows a susceptible - infectious pathway because of that resistance. We disregard the possibility of long-lasting immunity.

Susceptible individuals *S* can be infected by the pathogen variants through their interaction with the contaminated environment. If they are infected by an antimicrobial sensitive pathogen, an acute disease will be developed by a proportion *σ* of these hosts, transitioning to the normal infectious stage *I*. However, if susceptible individuals are infected by an AMR pathogen a proportion *ϵ* of them will transition to *C* showing more severe symptoms, and the other proportion (1 − *σ* − *ϵ*) will enter the compartment of asymptomatic carriers *A* until such a time when the disease becomes active (the host shows symptoms).

An important hypothesis in this study is that individuals classified as *A* are produced through the continuous exposure of naive susceptible individuals to contaminated environments [11, 20]. It suggests that the onset of an active (severe) antimicrobial-resistant (AMR) infection stems from ongoing, albeit random, interactions with the polluted environments where a mixture of sensitive and AMR pathogens are present. Moreover, we assume that asymptomatic individuals may progress to develop a recurrent disease due also to sustained exposure to contaminated environments, along with potential contributing factors in the host, such as comorbidities and malnutrition [22, 26].

Since the disease is transmitted through the environment, we assume that *I, C* and *A* individuals shed their pathogens into that environment at a rate *ξ* where all bacterial types completely mix. We also assume that there is an external contamination rate *ρ* that contributes to the bacterial level of the environment.

We set *γ* as the recovery rate for acute infections, causing the individuals in *I* to return to *S*. We also assume that individuals with severe disease can relapse into the asymptomatic compartment with rate *θ*, and subsequently move to the susceptible stage again with a rate *κ* [22, 33]. In general we assume that 1*/γ* is shorter that 1*/θ* because of the severity of the illness; also, we can assume *κ* is small since antimicrobial resistance would make it difficult to recover to a fully susceptible (healthy) stage.

Infection occurs when susceptible individuals get into contact with the polluted environment. The infection rate of the naive susceptible individuals *S*, is *β. A* individuals become symptomatically infected, i.e., develop an active infection, at a rate *ϕβ* where *ϕ* is a positive index 0 *< ϕ* indicating that hosts may have lower (0 *< ϕ <* 1) or higher tendency (1 *< ϕ*) to an asymptomatic infection.

The environment *E* is related to a measure of the level of pathogens in the environment but it comprises all sort of factors besides pathogens that may affect the susceptibility of a host to disease. We assume it presents a logistic dynamics with is growth rate given by the different shedding rates; the clearance rate is *ω*. In §5 the shedding rate *ξ* is weighted by the probability of disease progression to *I* or *C* forms. In what follows we consider the simple case where *ξ* is not weighted but is the same for all types of infections (see model below). Finally we assume that the host population size is constant.

The diagram of the model is shown in Figure 1 and the equations are:

**Figure 1.**
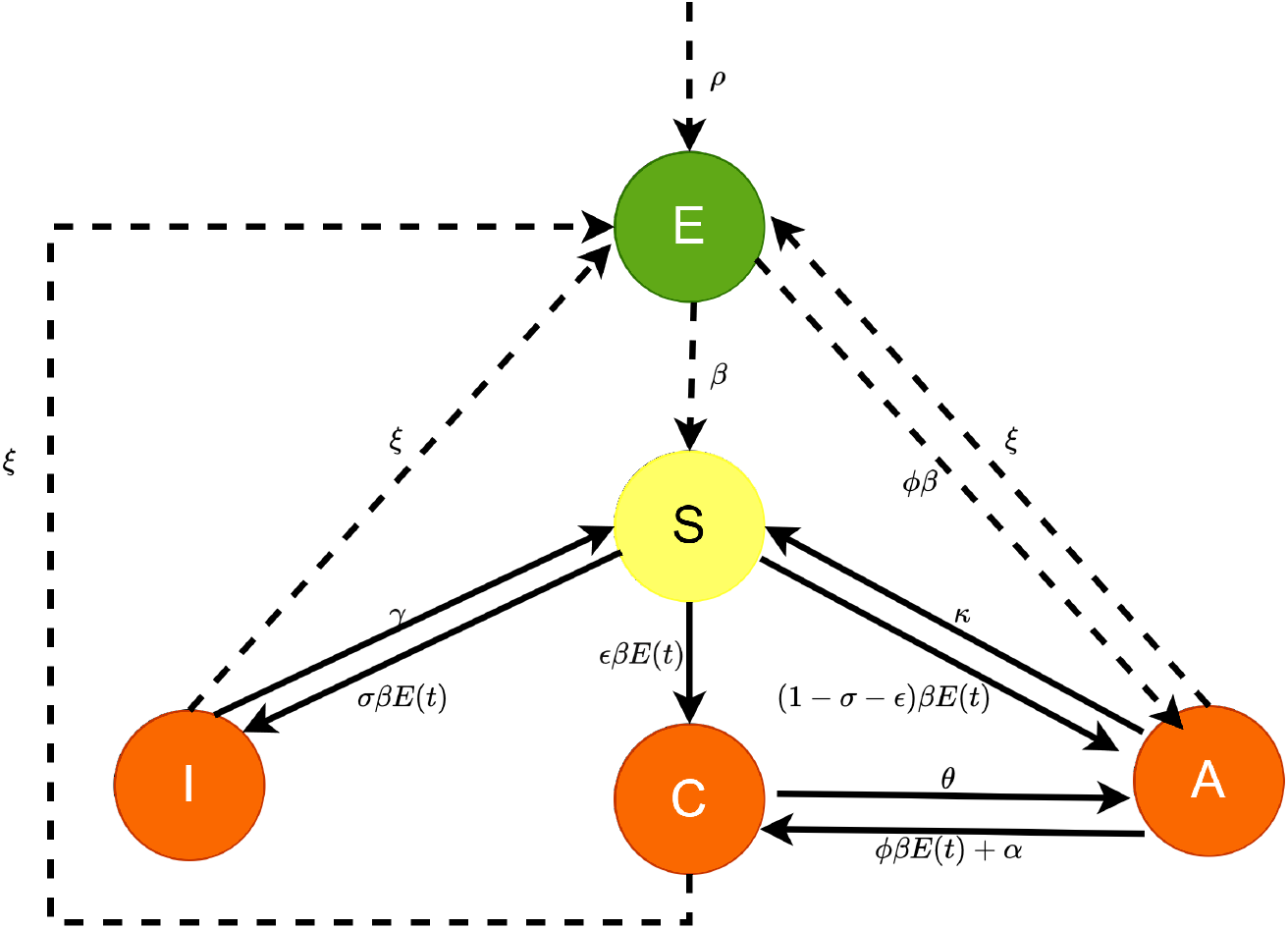
Diagram of model (1) where the environment *E* spreads two types of pathogens, one with antimicrobial resitance (AMR) and the other one with sensitive to them. Both of them can infect the susceptible population (S) developing an acute disease (I), a severe disease (C) or by becoming asymptomatic carries (A), who will recurrently develop a form of severe disease. On the other hand, the environment is a well mixed composite of all pathogen types. Also, we assume an external supply of contaminated environment *ρ*.

**Figure 2.**
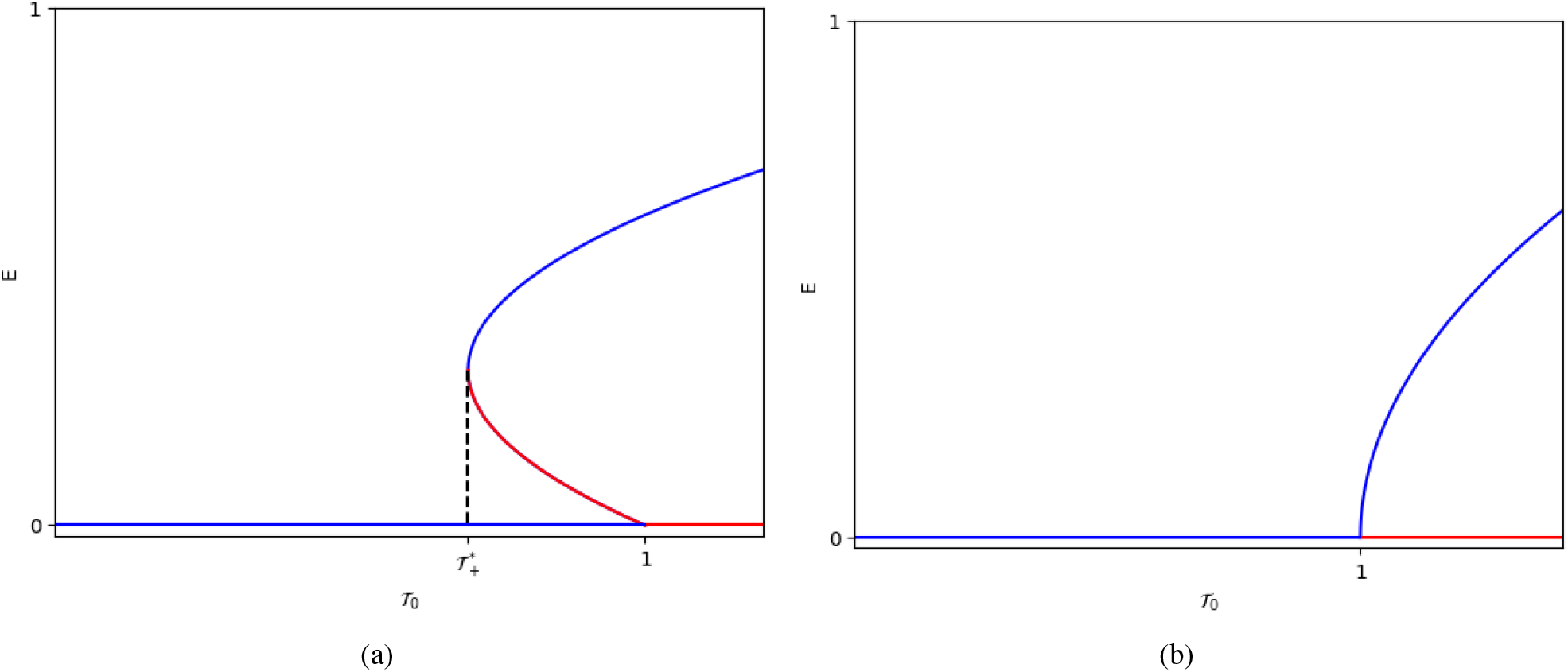
Bifurcation diagrams for system (2) in two scenarios: (a) ℛ_*_ *>* 1 (backward bifurcation); (b) ℛ_*_ *<* 1. Blue represents stable equilibria, and red represents unstable equilibria

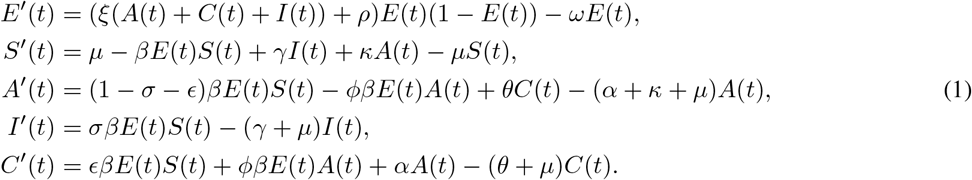

In order to simplify the calculations of the following analysis, we consider only the case *κ* = 0 and in §5 we will show that our results on the behavior of the system hold when considering *κ >* 0. Because of we assume a constant population with *S* + *A* + *I* + *C* = 1, it is possible to reduce the model (1) setting *C* = 1 − *S* − *A* − *I*, to obtain

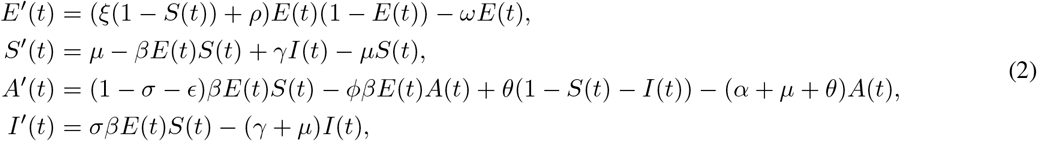

With the above, we establish the next proposition whose proof is omitted

### Proposition 1

*Let* Ω = {(*E, S, A, I*) | 0 ≤ *E*≤ 1, *S* ≥ 0, *A* ≥ 0, *I* ≥ 0, *S* + *I* + *A* 1,}. *Then* Ω *is a positively invariant set for (2)*.

## 3 Steady-states

### 3.1 Disease-free equilibrium

In order to find the disease-free equilibrium (DFE), we set *E* = 0, no polluted environment. Then *DFE* = (0, 1, 0, 0). To establish the local stability of this equilibrium, we compute the jacobian matrix of the system (2) evaluated at this point, obtaining

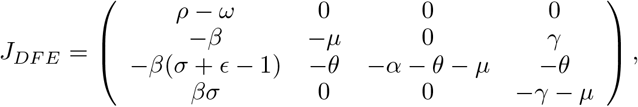

whose eigenvalues are {*ρ* − *ω*, −*μ*, −*γ* − *μ*, −*α* − *θ* − *μ*}. Therefore, the stability of *DFE* can only change through the eigenvalue *ρ* − *ω*, thus, we define an environmental threshold parameter as

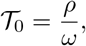

that depends only on the environmental pollution rates. Setting *ρ* = 𝒯_0_ *ω*, we can rewrite the eigenvalues of *J*_*DF E*_ as

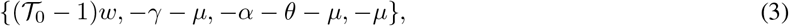

therefore, all of them are negative for 𝒯_0_ *<* 1 and we can establish the next proposition.

#### Proposition 2

*The disease-free equilibrium DFE is locally asimptotically stable for* 𝒯_0_ *<* 1 *and unstable for* 𝒯_0_ *>* 1.

### 3.2 Endemic equilibria

The endemic states of the system show interesting characteristics with respect to its dynamic properties. Recall that the environmental threshold parameter 𝒯 _0_ of our system, given in the previous section, depends only on the inflow and outflow rates of the contaminated environment. This curious result is further characterized here. First, let’s determine the endemic equilibria of (2) solving the system of equations

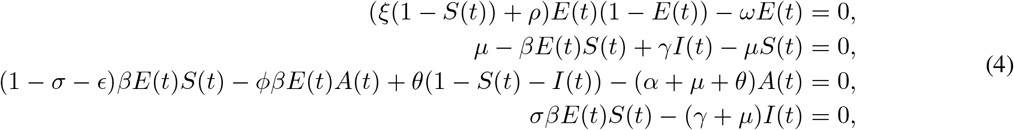

where we assume *E* ≥ 0. The equilibria are given by

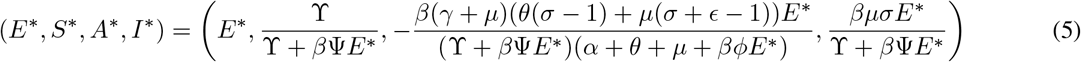

where we have defined two positive parameters, ϒ = *μ*(*μ* + *γ*) and Ψ = *μ* + *γ*(1 − *σ*), to simplify expressions, and *E*^*^ represents any root in the unit interval (0, 1), of the quadratic polynomial

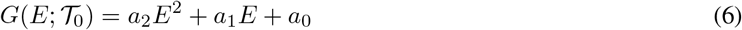

With

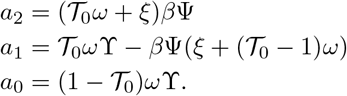

First of all, note that *a*_2_ *>* 0 always (since 𝒯_0_ *>* 0). Moreover, when 𝒯_0_ *>* 1, then *a*_0_ *<* 0 and

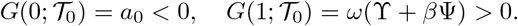

In this case, there exists exactly one endemic equilibrium point. However, if 𝒯 _0_ = 1, then *a*_0_ = 0 and there are two roots for *G* namely 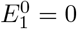, which coincides with the *DFE*, and a second root

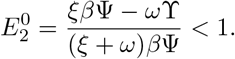

Note that if *ξβ*Ψ − *ω*ϒ *<* 0, then 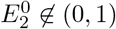, Thus, we establish the feasibility condition for this equilibrium

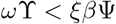

or, equivalently,

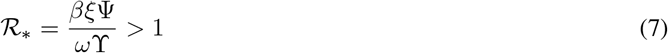

which can be given the following interpretation. Rewrite

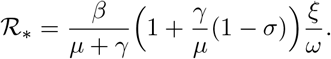

*β* is the *total* effective contact rate regardless of the type of infection that is produced; thus *β/*(*μ* + *γ*) is the total number of infections (regardless of its fate) that are produced during the average infectious period of an *I*-type individual; 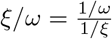 is the ratio of the life expectancy of the polluted environment to the duration of shedding. So, if we look at ℛ_*_ ℛ_*=_ (*σ*) as a function of *σ* we have that if *σ* = 1, meaning that all the incidence is directed to *I*-type individuals, then

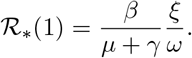

However, if no incidence is directed to the *I*-type individuals, i.e., *σ* = 0 then ℛ_*_ (0) *>* ℛ_*_ (1). Therefore, this threshold parameter describes the strength of the transmission to infectious classes other than the *I*-type infections. Note also that a fast *I*-type disease (1*/γ <* 1*/ω*), increases ℛ_*_for any *σ*.

Finally, it is easy to see that *G*(*E*; 𝒯_0_) is a continuously differentiable function and 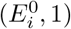 satisfies that 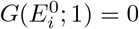, for *i* = {1, 2}, and

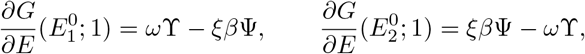

that is, 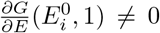.Then, by the Implicit Function Theorem [25], for each *i* there exists a unique function 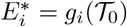 such that *G*(*g*_*i*_(𝒯_0_); 𝒯_0_) = 0 in a neighborhood of 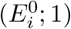, that is, these fuctions represent the roots of (2) in those neigborhoods. It is possible to write *g*_*i*_(𝒯_0_) as a series expansion around 𝒯_0_ = 1, obtaining

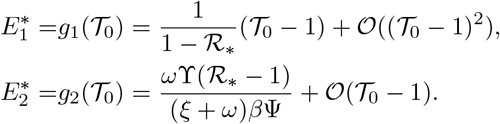

We can see that the condition ℛ_*_*>* 1 guarantees the existence of an open interval when 𝒯 _0_ *<* 1, such that the two roots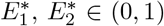, implying the existence of two endemic equilibrium points. In what follows we characterize these equilibria.

We first determine the threshold values for which the discriminant of the function *G*(*E*; 𝒯_0_) vanishes. These are

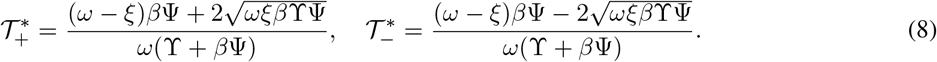

Each one of the above provides a unique solution for *G*(*E*; 𝒯_0_) = 0 given, respectively, by

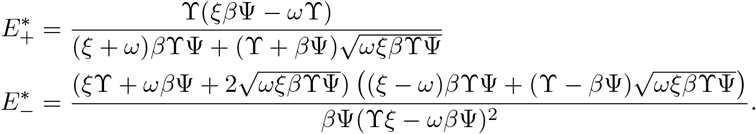

We now proceed to eliminate the pair 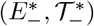 as an possible endemic equilibrium. Observe that 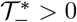. if and only if 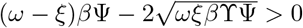 Besides, we have that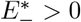 if and only if 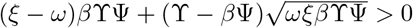 Thus, assume 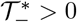, implying 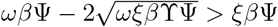, then

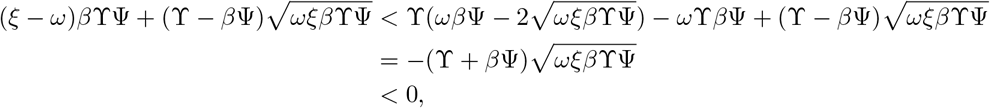

thus, the point with 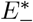 is not a biologically feasible endemic equilibrium. Now, for the pair 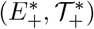, note that

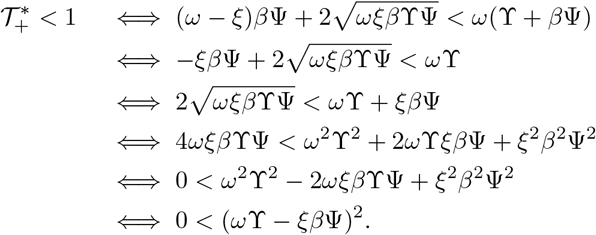

In other words, for this point 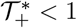 always. Finally by (7),

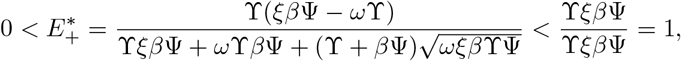

implying that the vertex of the parabola (6) lies on the unit interval (0, 1). We summarize the above analysis in the following

#### Proposition 3

*Consider system (2), with* 𝒯_0_ *and* 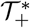 *defined above:*

i. *If* 𝒯_0_ *>* 1 *there exists a unique endemic equilibrium*.
ii. *If (7) is satisfied and* 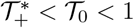 *then there are two endemic equilibria*.
iii. *Otherwise, there are no endemic equilibria*.

## 4 Bifurcation analysis

In this section we look closer to the bifurcation phenomenon described above using 𝒯_0_ as a parameter to obtain the bifurcation diagram for the equilibria. First, upon implicit differentiation of (6) with respect to 𝒯_0_ we obtain

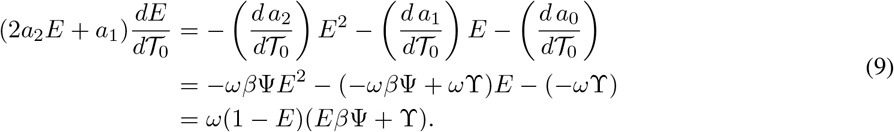

Clearly, the right-hand side of the above is positive, and therefore the sign of the slope of the bifurcation curve is determined by the sign of the term 2*a*_2_*E* + *a*_1_. If 𝒯_0_ *>* 1, the unique endemic equilibrium satisfies 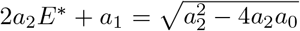, implying that the bifurcation curve has positive slope for *E*^*^ *>* 0. If, however, (7) is satisfied with 𝒯_0_ *<* 1, then there exists an interval where there are two branches of endemic equilibria: the larger one has positive slope 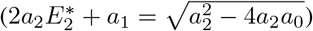 and the smaller one has negative slope 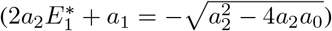.

For the stability of the endemic equilibria we look at the jacobian matrix of (2) evaluated at the equilibrium (*E*^*^, *S*^*^, *A*^*^, *I*^*^), obtaining

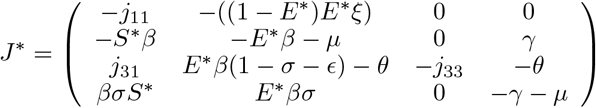

With

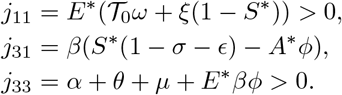

The trace *tr*(*J*^*^) *<* 0 and, using equation (4), the determinant is

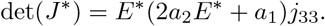

Also, the characteristic equation is

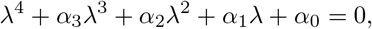

With

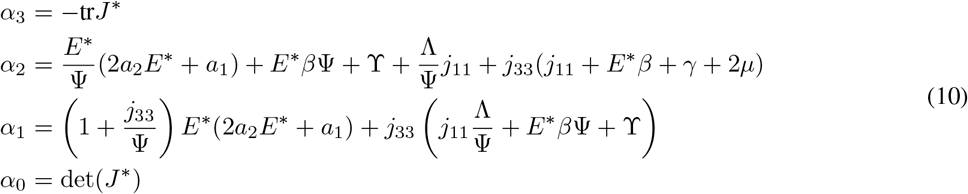

where Λ= *μ*Ψ + *γ*(*γ* + *μ*)(1 − *σ*) *>* 0. Moreover, we have

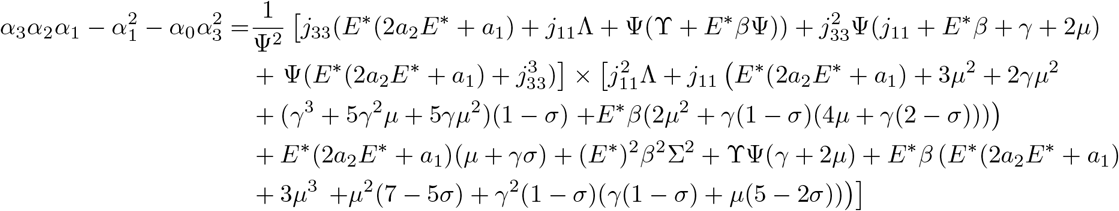

Clearly, if an endemic equilibrium branch has positive slope, we have *α*_*i*_ *>* 0 for *i* = 0, …, 3 and 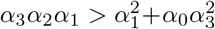 then, by the Routh-Hurwitz criterion [1], all the eigenvalues have negative real part. On the other hand, if an endemic equilibrium branch has negative slope this implies *α*_0_ *<* 0 and therefore some eigenvalues have non-negative real part. We establish the following result.

### Theorem 1

*If an endemic equilibrium of (2), is located on a bifurcation branch with positive slope, then it is locally asymptotically stable whereas if it is on a branch with negative slope, it is unstable*.

To further characterize this backward bifurcation, we will show now that two phenomena take place in system (2) when is satisfied: *a*) the DFE collapses onto the unstable endemic equilibrium generating a change of stability in the first one when *E*^*^ = 0 and 𝒯_0_ = 1; *b*) both bifurcation branches of endemic equilibria intersect rendering a single endemic equilibrium when 2*a*_2_*E*^*^ + *a*_1_ = 0, or equivalently, when 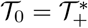; no endemic equilibria exist when 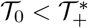.

First, let us define the function *F* (*E, S, A, I*; 𝒯_0_) = (*E*^′^(*t*), *S*^′^(*t*), *A*^′^(*t*), *I*^′^(*t*))^*T*^, and suppose that (7) is satisfied.

Next, as we can see from (5) and (6), the *DFE* collides with an endemic equilibrium when *E*^*^ = 0 and 𝒯 _0_ = 1, that is when *F* (0, 1, 0, 0; 1) = **0**. Moreover, we see from (3), that exactly one eigenvalue associated to this equilibrium is equal zero, and this eigenvalue has left and right eigenvectors given, respectively, by

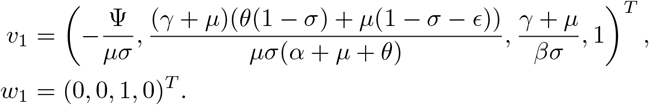

Finally, observe that 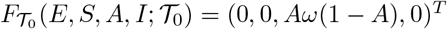, and from above results we obtain

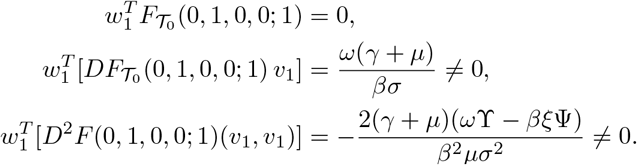

Thus, by Sotomayor theorem [29], system (2) presents a transcritical bifurcation at the *DFE*. We establish this result as follow.

### Theorem 2

*The system (2) experiences a transcritical bifurcation at the DFE as the parameter* 𝒯_0_ *passes through the bifurcation value* 𝒯_0_ = 1.

On the other hand, the determinant of the quadratic polynomial *G*(*E*; 𝒯_0_) vanishes when 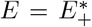 and 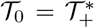, or, equivalently when 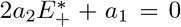. Let us define the endemic equilibrium corresponding to these values as 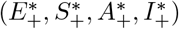 which satisfies 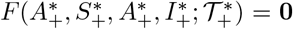. Also, from (10), we see that the Jacobian matrix at this point has exactly one eigenvalue equal zero and corresponding right and left eigenvectors, given by

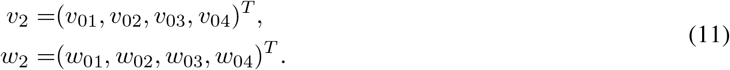

The explicit expressions of these eigenvectors are presented in A. With above results, we obtain

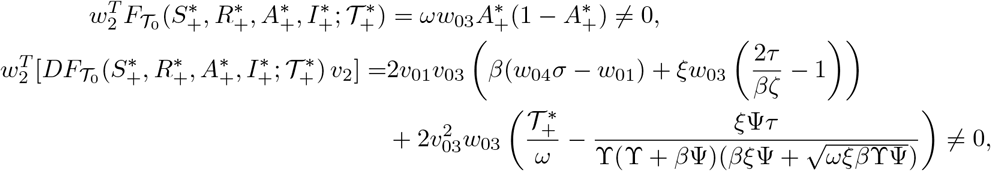

where *τ* and *ζ* are positive parameters defined, respectively, as

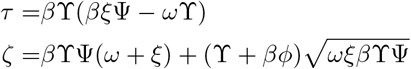

Thus, by Sotomayor theorem, system (2) presents a saddle-node bifurcation. We establish this result as follow.

### Theorem 3

*System (2) experiences a saddle-node bifurcation at* 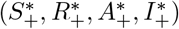 *as the parameter varies through the bifurcation value* 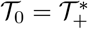.

In order to summarize all the analysis of this section, we rewrite the Proposition 3, characterizing the backward bifurcation.

### Theorem 4

*Consider the system (2), with* 𝒯_0_, 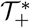 *and* ℛ_*_ *defined above:*

(i) *If* 𝒯_0_ *>* 1 *there exists a unique endemic equilibrium which is stable*.
(ii) *If* ℛ_*_ *>* 1 *and* 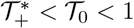 *then there are two endemic equilibria, one of them is stable and competes with the locally stable DFE*.
(iii) *Otherwise, the DFE is the unique atractor for* 𝒯_0_ *<* 1.

## 5 Numerical simulations

In this section we numerically explore the dynamic behaviour of model (2). We set the effective contact rate *β* = 0.9, the recovery rate of the acute illness *γ* = 1*/*6, and the recovery rate of severe illness *θ* = 1*/*30, the growth rate of the population is *μ* = 1*/*(70 × 365) and the infection rate of the asymptomatic and the susceptible individuals are equal, i.e., *ϕ* = 1; the rate of severe disease development is *α* = 1*/*100 and the shedding rate is *ξ* = 0.1.

Here, we exemplify the scenarios described in Theorem 4 by adjusting both, the external contamination rate, *ρ*, and the clearance rate *ω*. First, take *ρ* = 0.1 and *ω* = 0.16. These values render the environmental threshold parameters 𝒯_0_ = 0.625 and 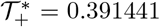 while the condition ℛ_*_*>* 1 is satisfied. In consequence, condition ii) of Theorem 4 guarantees that depending on initial conditions, we can obtain either that the infection reaches the disease free equilibrium point or the disease establishes at an stable endemic one.

**Scenario 1**. In Figure 3a we present the case for an 8% of contaminated environment and a completely susceptible population as initial conditions, *i*.*e*., *E*_0_ = 0.08, *S*_0_ = 1. In this case, the system reaches the endemic equilibrium point after one year where severe and asymptomatic compartments constitute 80% and 20% of population, respectively, and the contaminated environment reaches 20%. Notice, however, that at the beginning of simulation the level of contaminated environment actually diminishes to a value close to zero for most of the first year. after this transient, the asymptomatic disease begins to slowly increase, reaching around 40% of population in the first 200 days, this is followed by a decrease that finally set it around 20% of population. Note that the severe disease compartment grows throughout the whole time.

**Figure 3.**
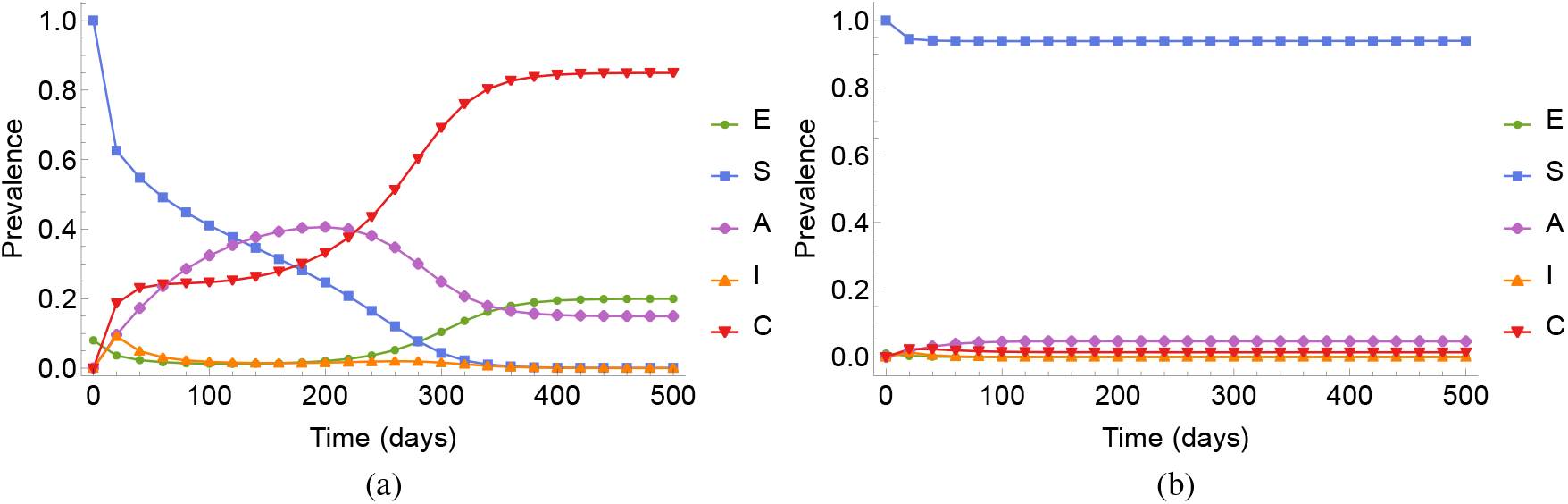
Simulations for system (2) with ℛ_*_*>* 1, 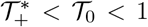, with *α* = 0.01, *β* = 0.9, *γ* = 1*/*6, *ϵ* = 0.3, *θ* = 1*/*30, *ξ* = 0.1, *σ* = 0.6, *ϕ* = 1, *ρ* = 0.1, *ω* = 0.16 (_0_ = 0.625), *μ* = 1*/*(70(365)). By Theorem 4, two locally stable equilibrium compete: (a) The endemic equilibrium point is reached with initial conditions *E*_0_ = 0.08, *S*_0_ = 1, *A*_0_ = 0 *I*_0_ = 0, *C*_0_ = 0. (b) The DFE point is reached with initial conditions *E*_0_ = 0.01, *S*_0_ = 1, *A*_0_ = 0, *I*_0_ = 0, *C*_0_ = 0.

**Scenario 2**. To illustrate other scenario, take *E*_0_ = 0.01; this gives a different behavior (see Figure 3b). The system reaches the DFE point, but during the first 500 days it appears converge to some endemic point with around 5% of asymptomatic infections. However, for longer times we observe a reduction of severe and asymptomatic cases, while a significant level of uncontaminated environment is maintained allowing thus the increase of the susceptible population.

**Scenario 3**. If we now set *ρ* = 0.1 and *ω* = 0.6 we still have that ℛ_*_ *>* 1 but ℛ_*0_ = 0.16 and 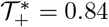are reduced which take us to the third condition of Theorem 4 that determines the approach of the system to the DFE (see Figure 4).

**Figure 4.**
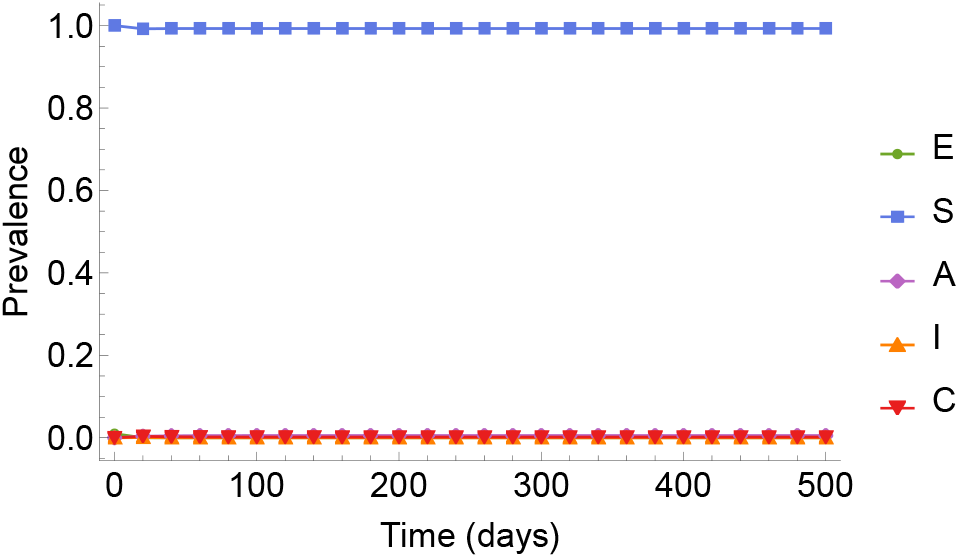
Simulations for system (2) with ℛ_*_*>* 1, 𝒯 _0_ *<* 1 but 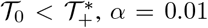, *α* = 0.01, *β* = 0.9, *γ* = 1*/*6, *ϵ* = 0.3, *θ* = 1*/*30, *ξ* = 0.1, *σ* = 0.6, *ϕ* = 1, *ρ* = 0.1, *ω* = 0.6 (𝒯 _0_ = 0.16) and *μ* = 1*/*(70(365)). By Theorem 4, the reach of the disease-free equilibrium point is assured with initial conditions *E*_0_ = 0.01, *S*_0_ = 1, *A*_0_ = 0 *I*_0_ = 0, *C*_0_ = 0.

**Scenario 4**. In contrast, Figure 5 shows the approach to the endemic equilibrium point when *ρ* = 0.12 and *ω* = 0.1. This implies that 𝒯_0_ *>* 1. It is necessary to emphasize that even when the contaminated environment shows a level lower than 60%, the population will always present signoficant asymptomatic and severe cases.

**Figure 5.**
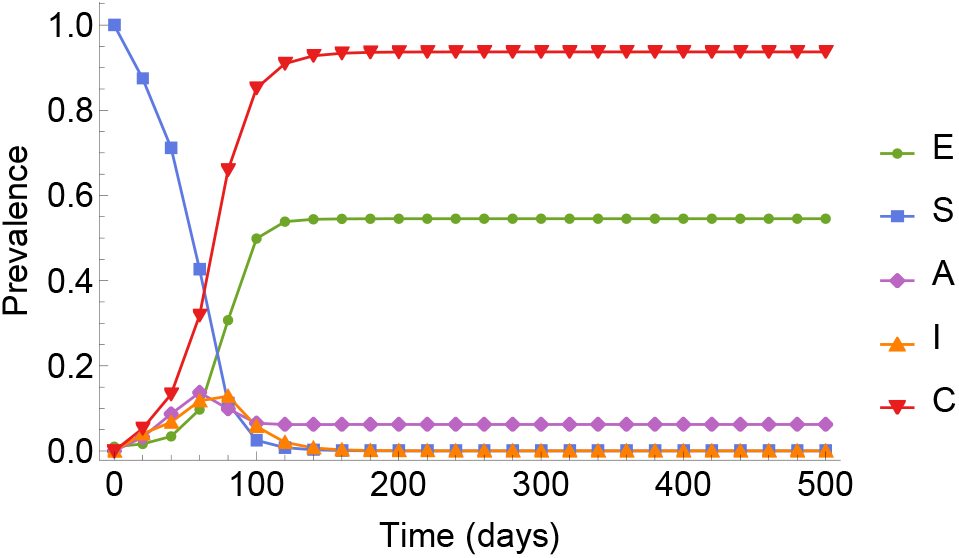
Simulations for system (2) with 𝒯 _0_ *>* 1 where the endemic equilibrium point is reached regardless of the initial condition established. Parameters values *α* = 0.01, *β* = 0.9, *γ* = 1*/*6, *ϵ* = 0.3, *θ* = 1*/*30, *ξ* = 0.1, *σ* = 0.6, *ϕ* = 1, *ρ* = 0.12, *ω* = 0.1 (𝒯 _0_ = 1.2) and *μ* = 1*/*(70(365)) and initial conditions *E*_0_ = 0.01, *S*_0_ = 1, *A*_0_ = 0, *I*_0_ = 0, *C*_0_ = 0.

**Scenario 5**. Finally, as mentioned in §2 the above simulations, as well as the previous mathematical analysis,concern the simplified mathematical model (2) when *κ* = 0. In this scenario we show that these results are robust for *κ >* 0, that is the backward bifurcation shown in Figure 2a is maintained for *κ* ∈ [0, 1] in the full model (1). Figure 6 shows the bifurcation diagram of (1) (with the parameters used in Figure 3a) for a range of values of *κ*. This suggests that even when a full recovery of asymptomatic individuals is possible and the environmental clearance rate is greater than its external contamination rate, there is bistability, i.e., under some initial conditions it is possible to achieve either the endemic equilibrium point or the disease free equilibrium point.

**Figure 6.**
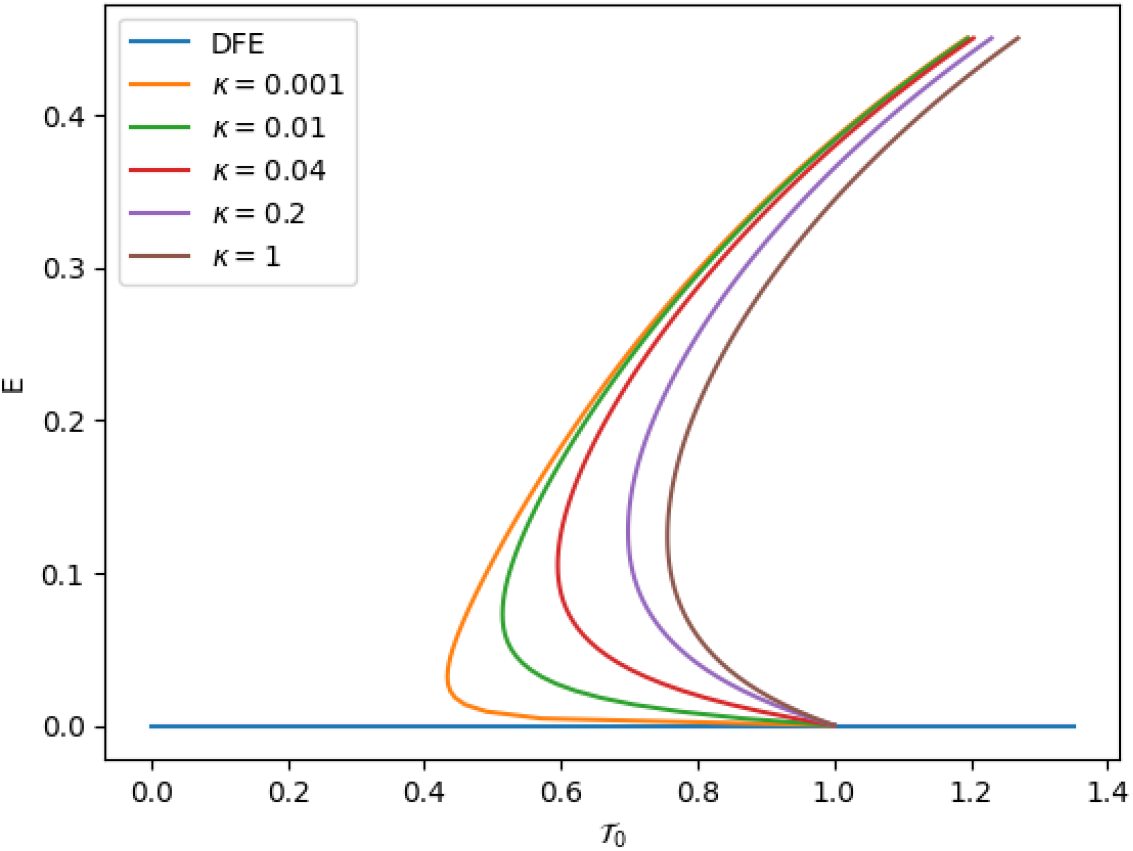
Bifurcation diagrams for system (1) considering some values for *κ*, and the values of the remaining parameters as in simulations of Figure 3a. The backward bifurcation is preserved for this values.

### 5.1 Generalizations

In some work related to environmental-transmission diseases, it is considered that the rate at which asymptomatic individuals shed their pathogens into the environment is lower than the rate at which those individuals who show symptoms do. To generalize our model to this case, we have to establish two rates of shedding, *ξ*_1_ for compartments *I* and *C* and *ξ*_2_ for *A*, with *ξ*_1_ *< ξ*_2_, setting the first equation of system (1) as

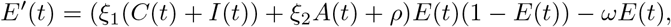

and after the substitution *C* = 1 − *S* − *I* − *A*, the first equation in (2) is

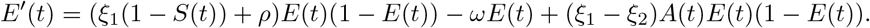

Let us refer to this system as (2a). We can see that system (2a) is positively invariant, has a DFE=(1,0,0,0) and the stability of this equilibrium is still given by 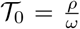, same as form system (2). The main difference between both systems is the difficulty of finding the endemic equilibria. *S*^*^,*A*^*^ and *I*^*^ and *E*^*^ where *E*^*^ is a root of

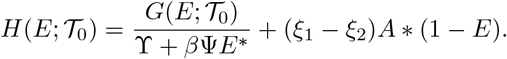

Note that when *ξ*_2_ = *ξ*_1_, then its roots are given by *G*(*E*; 𝒯 _0_). When different, the roots of *H*(*E*; 𝒯 _0_), are given by the cubic polynomial

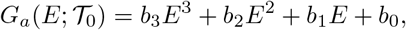

With

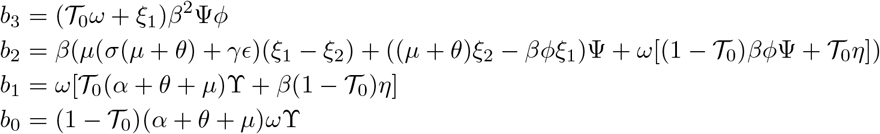

with the positive parameter *η* = (*α* + *θ* + *μ*)Ψ + *ϕ*ϒ. One of our future goals is to present a formal analysis for system (2a) similar to the one presented for system (2), where we expect to show a similar behavior. For example, if we use the parameters in Figure 3, setting *ξ*_1_ = *ξ* = 0.1 and *ξ*_2_ = 0.8, we obtain the polynomial

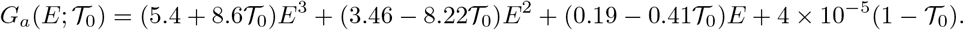

By the Descartes’ Rule of Signs, when 𝒯_0_ *>* 1, *G*_*a*_(*E*; 𝒯_0_) has exactly one positive root. For 𝒯_0_ = 1, the roots are 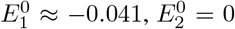, and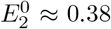. Finally, by the Implicit Function Theorem, the roots of *G*_*a*_(*E*; 𝒯_0_) around 𝒯_0_ = 1 are given by

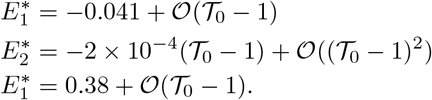

We can see that there exists an interval for 𝒯 _0_ *<* 1 where *G*_*a*_(*E*; 𝒯 _0_) has two positive roots. Then, the system (2a) has the same bifurcation diagram than system (2).

## 6 Conclusions

We have constructed, analyzed and interpreted a mathematical model for an environmental transmitted disease characterized for the existence of three disease stages. The pathways of the disease are illustrated in Figure 1. There are several key features in the model. First, *A*-type asymptomatic carrier individuals may be associated with AMR infections, and can arise by continuous exposure to a well mixed contaminated environment which is host to several strains of the pathogen. These *A*-type individuals, when continuously exposed to the contaminated environment, give rise to an active infection form of the disease and may reverse to asymptomatic carriers after some time i.e., become *A*-type individuals again. Second, *S*-type susceptible individuals, when infected, can follow the normal illness pathway or can become infected with what we call the acute form of the disease as in cholera, tuberculosis, toxoplasmosis and Giardia [15, 26, 27, 35]. Our main result is that the model equilibria may arise through a normal transcritical (forward) bifurcation depending only on the net increase in the level of environmental contamination determined by the threshold environmental parameter 𝒯_0_ = *ρ/ω*, which is independent of all transmission parameters; surprisingly, it only depends on those describing the input and output flows of the contaminated environment. However, disease dynamics driven by the contact rates (as normally occur in directly-transmitted or vector-transmitted diseases) only happens when the parameter *R*_*_ > 1 and, in this case, the forward transcritical bifurcation becomes a backward bifurcation, producing multiple steady-states, a hysteresis effect and dependence on initial conditions. This type of phenomenon has been described before in environmental diseases [14, 13]. The environmental parameters, still appear in this reproduction number, but they are represented through the ratio of pathogen shedding to the environmental clearance rate, *ξ/ω*, so shedding substitutes the input flow *ρ*. As described above ℛ_*_is a threshold parameter that describes the strength of the transmission to infectious classes other than the *I*-type infections. Also a fast *I*-type disease (1*/γ <* 1*/ω*), increases ℛ_*_ for any *σ*, the susceptibility index of *I*-type individuals.

## Data Availability

All data produced in the present work are contained in the manuscript

## Acknowledgements

We acknowledge support from DGAPA-PAPIIT-UNAM grant IV100220.

## A Eigenvectors for saddle-node bifurcation

In §4 we have used the eigenvectors (11), but, for simplicity, we have not presented the values of their components. In this section we provide these values. For *v*_2_ = {*v*_01_, *v*_02_, *v*_03_, *v*_04_}:

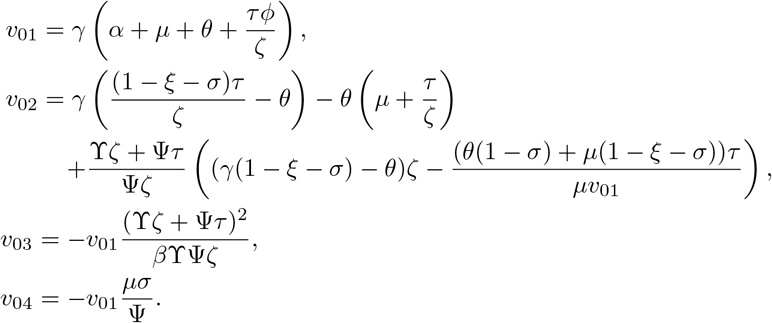

For *w*_2_ = {*w*_01_, *w*_02_, *w*_03_, *w*_04_}:

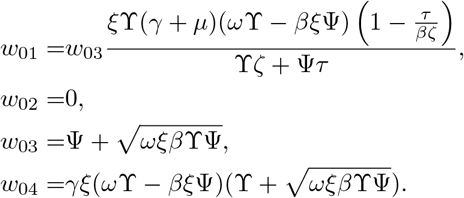

